# Impact of Nutrition, Sleep and Physical Activity on Cognitive Function in Older Frailty Adults: A Randomized Controlled Trial

**DOI:** 10.1101/2025.03.14.25323203

**Authors:** Keisuke Sakurai, Izumi Shiraishi, Hiroyasu Kasahara, Saya Anzai, Ryo Tanaka, Miwa Watanabe, Momoko Funakawa, Saori Shimada, Tatsuya Goto, Riki Kawabata, Keiko Murasakino, Tomoko Kawaura, Noriko Inamura, Ko Kaneta, Satoru Kobayashi, Toshiyuki Nomura, Nori Karasawa, Takashi Matsumoto, Tomoki Taira, Toshihiro Ishiguro, Kazuyuki Kudo, Yukihiro Sugawara, Hiroki Kayama, Matthew J. Thompson, Joseph Ledsam, Shin’Ichi Warisawa, Tatsuhiro Hisatsune

## Abstract

**Background:** As the global population ages, there is an increasing demand for effective strategies to maintain and improve health among older adults. Wearable technology presents a promising tool for health monitoring and management, yet its effectiveness in comprehensive health improvement for older adults remains uncertain.

**Objective:** This study aimed to evaluate the effectiveness of personalized lifestyle notifications, based on wearable device recorded data, in improving health outcomes, specifically cognitive and physical function, among older adults, compared to usual care.

**Methods:** In a 6-month randomized controlled trial, 355 older adults (aged 65+), including those with frailty, were randomly assigned to an intervention group (*n*=178) or a control group (*n*=177). The intervention group wore Fitbit Charge 5 devices and received personalized lifestyle alerts with rule-based personalization, using thresholds derived by human experts, throughout the 6-month period. The control group received no such notifications and were instructed not to use wearable devices. Some opt-in subjects, an intervention group (*n*=128) or a control group (*n*=116), were requested to record all meals using the application to deliver nutritional alerts. Cognitive function was assessed using the Montreal Cognitive Assessment (MoCA). Physical function was evaluated using Fried Frailty Phenotype criteria. Measurements were conducted at baseline and after 6 months.

**Results:** In the intention-to-treat analysis, the intervention group showed significant improvements in general cognitive function (MoCA scores increased by 1.0 (95% CI: 0.6 to 1.3) vs 0.2 (−0.2 to 0.5) in the control group, *p*=0.011) and frailty status (Fried frailty phenotype index change: −0.3 (−0.5 to −0.2) vs −0.1 (−0.2 to 0.1) in the control group, *p*=0.029). Subgroup analysis of participants with nutritional tracking showed significant improvements in MoCA scores (1.2 (0.8 to 1.6) vs 0.2 (−0.2 to 0.5), *p*=0.0004) and frailty status (−0.3 (−0.5 to −0.2) vs 0.0 (−0.2 to 0.1) in the control group, *p*=0.009). The per-protocol analysis showed similar results.

**Conclusion:** This study provides evidence that personalized, multifaceted Fitbit-based interventions can effectively enhance cognitive function, with notable improvements specifically in MoCA scores, and mitigate frailty progression in older adults as expected. These findings suggest that comprehensive lifestyle interventions including exercise, sleep and nutrition using wearable technology may be valuable for promoting healthy aging.

## 1. Introduction

As the global population ages, there is an increasing need for effective strategies to maintain and improve health among older adults [World Health Organization, 2015]. The health of older adults is multifaceted, with cognitive and physical functions being crucial components. While cognitive decline is a part of normal aging, its progression varies significantly among individuals and can potentially be improved or maintained through appropriate interventions [Murman, 2015]. Similarly, frailty, a syndrome that significantly impacts the independence and quality of life of older adults, requires early intervention [Fried *et al*., 2001; Hoogendijk *et al*., 2019].

In recent years, wearable technology has emerged as a promising tool for health monitoring and management. These devices continuously collect health-related data such as physical activity, sleep patterns, and heart rate, providing users with real-time feedback. Intervention studies utilizing activity trackers have reported improvements in health behaviors, particularly increased physical activity [Brickwood *et al*., 2019].

While the potential benefits of wearable technology are clear, its use among older adults presents unique challenges, such as technology acceptance and long-term motivation [Kononova *et al*., 2019]. Research has shown that older adults tend to exhibit more positive attitudes and a higher sense of activity obligation when using activity monitoring technology compared to their younger counterparts [Hamido *et al*., 2021]. This suggests that, despite potential hurdles, wearable technology may hold particular promise for improving health outcomes in older populations.

Personalized lifestyle interventions, tailored to individual needs and characteristics, have shown promise in promoting behavior change and health improvements [Michie *et al*., 2017]. Individualized notifications and advice based on data from wearable devices may enhance adherence to health-promoting behaviors and lead to improved health outcomes. Recent research has highlighted the complex interplay between cognitive frailty and physical health conditions, such as heart failure [Yamamoto *et al*., 2022], underscoring the importance of a holistic approach to health interventions for older adults.

Previous research has demonstrated that multidomain interventions combining diet, exercise, cognitive training, and vascular risk management may prevent cognitive decline in older adults [Kivipelto *et al*., 2020; Rosenberg *et al*., 2018]. However, the impact of comprehensive interventions using wearable technology on both cognitive function and frailty has not been thoroughly investigated.

To address this gap, we investigated the effect of a complex intervention of lifestyle notifications based on wearable device-recorded data on cognitive function and physical health outcomes among older adults, compared to usual care.

## 2. Materials and Methods

### 2.1 Study Design

The goal of this prospective, single site, interventional randomized control trial is to evaluate whether the use of Fitbit devices improves functional independence in adults aged 65 years and older. The main question of the study is to determine the effectiveness of notification on activity, sleep, and nutrition based on wearable device Fitbit recorded data for the improvement of health conditions including intellectual property and physical function, compared to usual care. Participants in the intervention arm were given a Fitbit Charge 5 device and asked to wear this for the duration of the study. The study intervention ran for 6 months. Notifications were issued using monitoring software, and issued automatically to participants.

### 2.2 Participants

We recruited 355 community-dwelling adults aged 65-93 years. The study population was drawn from the larger Kashiwa Study Cohort, a prospective cohort study designed to characterize biological, psychosocial, and functional changes associated with aging in a community-based sample of 2044 older adults living in Japan [Suthutvoravut *et al*., 2019]. The recruitment process involved sending study announcements with enclosed reply postcards by mail to eligible individuals registered in the Kashiwa Study Cohort. Those who returned the postcard indicating their interest were contacted via telephone to schedule an initial visit for informed consent discussion and screening.

Inclusion criteria were: age 65 years or older and less than 93 years, living independently with or without carer support, ability and willingness to wear a Fitbit device for the duration of the study (≥20 hours a day, including during sleep), smartphone access to allow Fitbit installation and setup, capacity to consent to study inclusion, and presence of either comorbidities or physical decline indicators. Comorbidities included low Body Mass Index (BMI) (<20 for ages 65-69, <21.5 for ages ≥70). Physical decline was defined as pre-frailty or frailty (scoring 1-2 or ≥3 respectively on the Fried Frailty Phenotype criteria [Fried et al., 2001]), or sarcopenia defined by low grip strength (<18 kg for female, <28 kg for male), slow gait speed (<1 m/s), or low skeletal muscle mass index (SMI) (<5.7 kg/m² for female, <7 kg/m² for male).

Exclusion criteria encompassed full-time care facility residence, inability to receive and/or act on interventions, prior experience using wearables to manage health, uncontrolled hypertension, diabetes, or chronic kidney disease. Additionally, individuals with cardiac pacemakers and/or cardiovascular stents were excluded. The Principal Investigator or designee reserved the right to exclude any individual deemed inappropriate for participation due to any condition or situation. Furthermore, individuals without any of the specified comorbidities were not eligible for inclusion.

During the screening process, potential participants underwent a comprehensive evaluation to ensure they met all inclusion criteria and did not fall under any exclusion criteria. If a participant met all requirements and expressed continued interest in the study, written informed consent was obtained using the institutional review board approved informed consent form. Participation was completely voluntary, and each participant was free to withdraw from the study at any time.

### 2.3 Randomization and Blinding

Participants were randomly assigned to the intervention (*n*=178) or control (*n*=177) group using block randomization in blocks of four, using a pre-generated randomization schedule. Due to the nature of the intervention, blinding was not possible for participants or the research team administering the intervention. To mitigate potential bias and any Hawthorne effect, we implemented several strategies. Upon completion of the study, all participants received a Fitbit device as compensation for their full participation. This approach helped maintain similar levels of engagement and attention across both intervention and control groups. Additionally, investigators were blinded to randomization during outcomes assessment. We employed a partial blinding technique by informing all participants that they were part of a study examining the effects of wearable devices on health outcomes, without specifying the exact nature of the intervention alert notification. This approach helped minimize expectation bias and reduced the potential for differential behavior change based on group assignment.

### 2.4 Device Description

Participants used Fitbit Charge 5 devices. The Fitbit Charge 5 device was first released in 2021. It is a slim fitness tracker with built in GPS, that includes sensors to track activity, sleep, SpO2, heart rate, skin temperature, breathing rate and stress. The device itself stores 7 days of detailed motion data, minute by minute and daily totals for the last 30 days. Data is stored in the corresponding Fitbit app, and is uploaded via Bluetooth when the app is paired to the wearable device. The Charge 5 housing is made of aluminum, glass and resin. The band is made of silicone and has an aluminum buckle, the device measures 36.7mm * 22.7mm * 11.2mm (not including the strap) and weighs 28g. The Charge is water resistant to 50 meters. It is compatible with Apple iOS 14 or higher and Android OS 8 or higher. If a device deficiency occurred, participants were asked to bring the device to the study site for a replacement device.

### 2.5 Intervention

Participants in the intervention group were provided with a Fitbit Charge 5 device and were asked to continuously throughout the study period. Participants were instructed to charge their device every 2-3 days, preferably during bath time or other activities when they would typically remove the device, to ensure continuous activity data collection throughout their active hours. The device’s battery life is approximately 7 days. The device collected data on physical activity (step count and active minutes) and sleep patterns (duration and onset time). Data were analyzed in both monthly and weekly intervals, with average daily values calculated to account for day-to-day variations.

#### 2.5.1. Automated Alert System

An automated alert system sent personalized text messages (**Table 1**) to participants on both monthly and weekly bases based on their activity and sleep data. The monthly alerts categorized behaviors into four levels: insufficient, somewhat insufficient, sufficient, and excellent for step count and active minutes, with an additional ‘excessive’ category for sleep duration. For step count, less than 3,000 steps per day was considered insufficient, 3,000-5,999 somewhat insufficient, 6,000-7,999 sufficient, and 8,000 or more excellent. Active minutes were categorized as insufficient if less than 45 minutes per day, somewhat insufficient for 45-89 minutes, sufficient for 90-149 minutes, and excellent for 150 minutes or more. Sleep duration was deemed insufficient if less than 5 hours per night, somewhat insufficient for 5-5.9 hours, sufficient for 6-8.9 hours, and excessive for 9 hours or more.

Weekly alerts were designed to provide more frequent feedback on participants’ progress. For step count and exercise time, these alerts included congratulatory messages for maintaining excellent levels (mean daily≥8,000 steps or ≥150 minutes) or achieving ≥10% improvement from the previous two weeks. Additionally, sleep-related weekly alerts monitored not only duration but also the variability in bedtime and wake times (**Table 1**), with alerts triggered when the average variability exceeded 30 minutes.

**Table 1:** Intervention Alert Logic Based on Fitbit Data.

| Alert Type | Criteria | Message Content | Goal |
| --- | --- | --- | --- |
| Monthly Step Count Alert | Average daily steps < 3000 | Encouragement to increase steps | Average daily steps ≥ 3000 |
| | $3000 \leq$ Average daily steps < 6000 | Encouragement to increase steps | Average daily steps ≥ 6000 |
| | $6000 \leq$ Average daily steps < 8000 | Encouragement to increase steps | Average daily steps ≥ 8000 |
| | Average daily steps ≥ 8000 | Congratulatory and encouragement to maintain steps message | Average daily steps ≥ 8000 |
| Monthly Exercise Time Alert | Average daily exercise time < 45 min | Encouragement to increase exercise time | Average daily exercise time ≥ 45 min |
| | $45 \text{ min} \leq$ Average daily exercise time < 90 min | Encouragement to increase exercise time | Average daily exercise time ≥ 90 min |
| | $90 \text{ min} \leq$ Average daily exercise time < 150 min | Encouragement to increase exercise time | Average daily exercise time ≥ 150 min |
| | Average daily exercise time ≥ 150 min | Congratulatory and encouragement to maintain exercise time message | Average daily exercise time ≥ 150 min |
| Monthly Sleep Duration Alert | Average daily sleep duration < 5 hours | Encouragement to increase sleep duration | Average daily sleep duration ≥ 5 hours |
| | $5 \leq$ Average daily sleep duration < 6 hours | Encouragement to increase sleep duration | Average daily sleep duration ≥ 6 hours |
| | $6 \leq$ Average daily sleep duration < 9 hours | Congratulatory and encouragement to maintain sleep duration message | $6 \leq$ Average daily sleep duration < 9 hours |
| | Average daily sleep duration ≥ 9 hours | Oversleep caution | $6 \leq$ Average daily sleep duration < 9 hours |
| Weekly Step Count Alert | Average daily steps ≥ 8000 | Congratulatory and encouragement to maintain steps message | Average daily step count ≥ 8000 |
| | Average daily steps < 8000 and step count increased by ≥ 10% compared to two weeks prior | Congratulatory and encouragement to improve steps message | Average daily step count increased by ≥ 10% compared to two weeks prior |
| | Average daily steps < 8000 and step count < 10% increase | Encouragement to improve | Average daily step count increased by ≥ 10% compared to two weeks prior |
| Weekly Exercise Time Alert | Average daily exercise time ≥ 150 min | Congratulatory and encouragement to maintain exercise time message | Average daily exercise time ≥ 150 min |
| | Average daily exercise time < 150 min and exercise time increased by ≥ 10% compared to prior week | Congratulatory and encouragement to improve exercise time message | Average daily exercise time increased by ≥ 10% compared to two weeks prior |
| | Average daily exercise time < 150 min and exercise time < 10% increase | Encouragement to improve | Average daily exercise time increased by ≥ 10% compared to two weeks prior |
| Weekly Sleep Duration Alert | Average sleep < 5 hours | Encouragement to increase sleep duration | Average daily sleep duration ≥ 5 hours |
| | $5 \leq$ Average sleep < 6 hours | Encouragement to increase sleep duration | Average daily sleep duration ≥ 6 hours |
| | $6 \leq \text{Average sleep} < 9 \text{ hours}$ | Congratulatory and encouragement to maintain sleep duration message | $6 \leq \text{Average daily sleep} < 9 \text{ hours}$ |
| | $\text{Average sleep} \geq 9 \text{ hours}$ | Oversleep caution | $6 \leq \text{Average daily sleep} < 9 \text{ hours}$ |
| Weekly Sleep and Wake Variability Alert | Average bedtime or wake-time variability $< 30 \text{ min}$ | Congratulatory and encouragement to maintain sleep and wake variability message | Average bedtime or wake-time variability $< 30 \text{ min}$ |
| | Average bedtime or wake-time variability $\geq 30 \text{ min}$ | Encouragement to improve | Average bedtime or wake-time variability $< 30 \text{ min}$ |

This table outlines the intervention alert logic used in the Fitbit-based data, detailing criteria for step count, exercise time, sleep duration, and sleep/wake variability. Data is collected via Fitbit devices and processed monthly and weekly to determine appropriate alerts, which are sent as SMS messages to participants. Each category of alerts is defined as follows: Alert Type: The specific health metric monitored and frequency. Criteria: The conditions based on Fitbit data that trigger alerts. Monthly alerts focus on average daily measures, while weekly alerts compare metrics to previous data (e.g., two weeks prior for steps). Message Content: Sample content of the SMS message sent to participants, which may include motivational messages, congratulations, or suggestions for improvement.

Alerts encouraged continuous improvement, prompting participants to aim for the next higher category or maintain excellent behaviors. For sleep, both insufficient and excessive durations were addressed to guide participants towards the sufficient range. Active minutes were calculated using Fitbit’s Active Zone Minutes system (https://help.fitbit.com/articles/en_US/Help_article/1379.htm), based on heart rate data. **Table 1** provides detailed information on alert content and logic for generating personalized recommendations.

The control group participants did not use Fitbit devices during the intervention period, nor did they receive personalized interventions. They were however provided with Fitbit devices and instructed to wear them only during two designated periods: the first and last two weeks of the study. This selective wearing schedule allowed us to collect comparable baseline data for both groups. To ensure protocol adherence, we sent email reminders to participants about the proper timing of device usage. During non-measurement periods, participants stored their devices at home.

#### 2.5.2 Nutritional Tracking

For nutritional tracking, participants who expressed willingness to keep food records at the start of the study were included in this aspect of the intervention. These participants used a food management app: *Asken* (see www.askendiet.com) to record their daily food intake.

Protein intake was categorized based on the percentage of total energy expenditure into four levels. This categorization referenced the approach used in the Kawasaki Aging and Wellbeing Project [Kurata *et al*., 2023] and was adapted to align with our study’s specific nutritional targets: insufficient (<13%), somewhat insufficient (13-14.9%), sufficient (15-19.9%), and excessive (≥20%). The messages encouraged participants to either achieve the next higher category or maintain sufficient intake levels, with specific dietary recommendations tailored to their current status (**Table 2**). These notifications formed the core of the nutritional intervention, providing guidance that participants could apply flexibly based on their dietary preferences and circumstances.

**Table 2:**
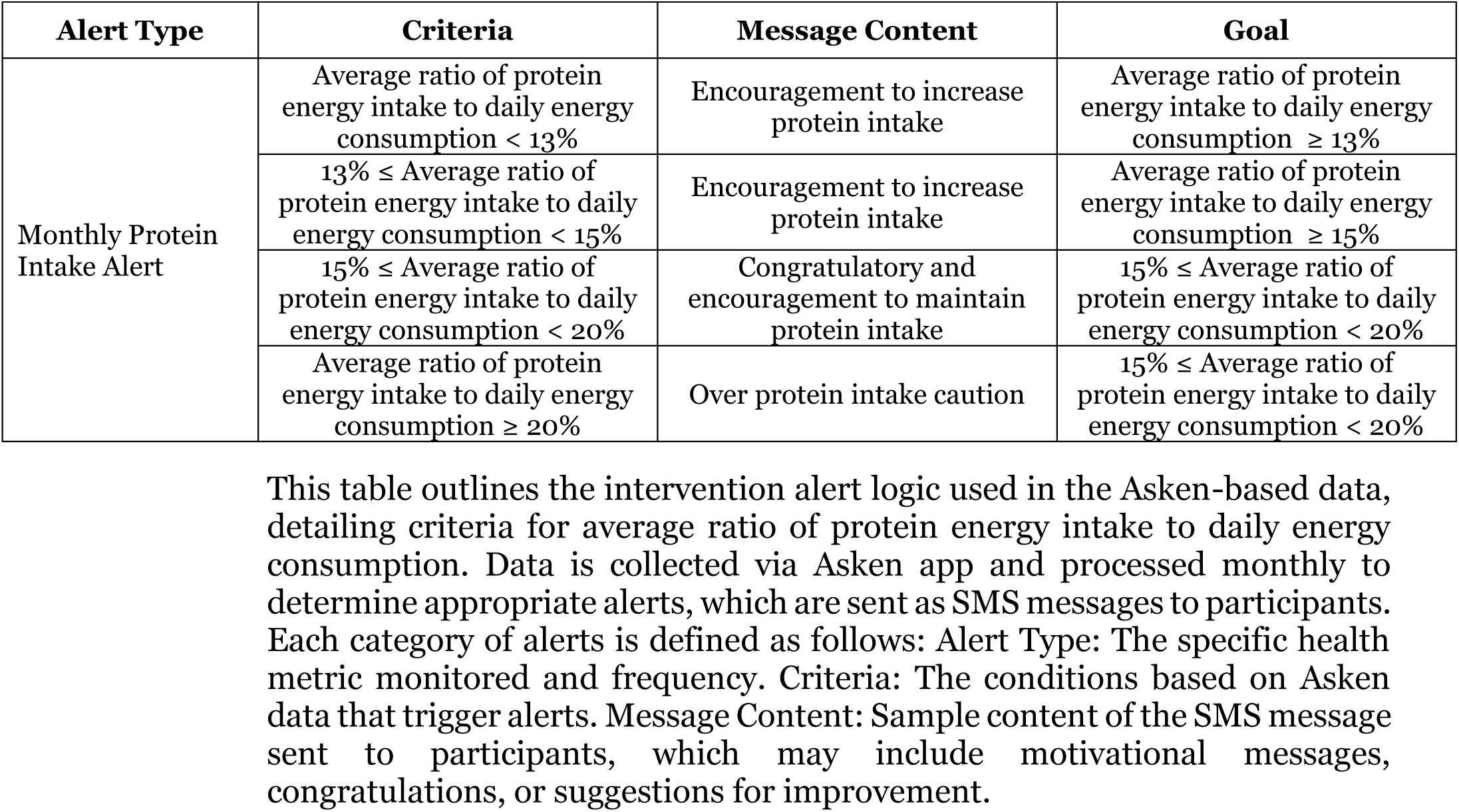
Comprehensive Nutritional Intervention Alert Logic Based on Asken Data.

Recognizing the potential financial barriers to increasing protein intake, we offered an optional supplementary food. Participants in the intervention group who expressed interest were provided with protein balanced food products free of charge. This option was designed to support those who might face economic constraints, offering a practical resource without prescribing it as a necessary component of the intervention. These foods were specially formulated using chicken and soy protein to provide 16.2g of protein per recommended daily serving, with a balanced ratio of 50% animal protein and 50% plant protein. This composition addresses both nutritional adequacy and sustainability concerns, while utilizing protein sources commonly consumed in the Japanese diet. The spherical shape was chosen to maximize versatility in cooking applications, allowing the product to be easily added to soups, stir-fries, or consumed as a standalone protein source. The protein balanced foods were manufactured by NH Foods Ltd (**Supplementary Table 1**). In the control group, only participants who expressed willingness to keep a food record at the beginning of the study were tracked for nutrition using Asken, but no intervention alerts or foods were provided.

### 2.6 Measurements

All outcomes were measured at baseline and 6-month follow-up. The baseline measurement for the Trail Making Test (TMT) was repeated on a different day due to a procedural error, and the conditions differed between the baseline and follow-up assessments. The assessment tools were selected because they showed validity and reliability in elderly populations across multiple countries.

General cognitive function was evaluated using the Montreal Cognitive Assessment (MoCA), which has demonstrated sensitivity to mild cognitive impairment [Nasreddine *et al*., 2005]. Executive function was assessed using the TMT Parts A and B (TMT-A/B), which measure components of executive control including visual attention, task-switching ability, and processing speed [Tombaugh, 2004]. Due to differing conditions during baseline and follow-up testing, the TMT results are reported in the supplementary materials as they may have introduced bias.

Physical status was evaluated through multiple measures. Muscle mass was quantified using bioelectrical impedance analysis. Physical frailty was assessed using the Fried Frailty criteria, comprising five components: handgrip strength (measured by dynamometer), walking speed (measured over a 5-meter course), self-reported unintentional weight loss, self-reported exhaustion, and self-reported physical activity level. Participants were classified as frail (three or more criteria met), pre-frail (one to two criteria), or non-frail (zero criteria).

Physical activity and sleep parameters were measured using Fitbit devices. Activity metrics included daily step counts, caloric expenditure, active minutes (categorized as fairly, lightly, or very active), and sedentary time. Sleep parameters encompassed total sleep time and sleep stages (light, REM, deep sleep, and awake in bed time). The System Usability Scale (SUS) [Brooke, 1996] was used to evaluate participants’ satisfaction with and usability of the Fitbit device and intervention program.

Nutritional status was monitored using the Asken food management application, which has been validated for Japanese populations, with particular emphasis on protein intake assessment. Adverse health events, including cardiovascular events (myocardial infarction, stroke), falls, exacerbation of chronic conditions, and acute hospitalizations, were also tracked as outcome measures.

### 2.7 Data Collection

Trained research staff conducted all clinical measurements at the study site following standardized procedures to ensure consistency across both study groups. Fitbit data were accessed through the Fitbit application programming interface (API) by authorized research staff for endpoint analysis and intervention notification alerts generation. Dietary information was collected through the Asken API, providing detailed nutritional intake data.

Semi-structured interviews were conducted with intervention group participants after the completion of all interventions and measurements to gather qualitative feedback about their experience. Participant questionnaires were administered to collect subjective data on health-related quality of life for all participants, and the System Usability Scale for the intervention group. Results of qualitative analyses will be presented in a subsequent manuscript.

To maintain data security and participant confidentiality, all study records and documents, including signed consent forms, will be retained by the Principal Investigator for 5 years after study completion. All data collection procedures were standardized to ensure reliability across both study groups.

### 2.8 Sample Size Calculation

The study aimed to recruit a total of 350 participants, with 175 individuals allocated to each of the intervention and control groups. Accounting for an anticipated dropout rate of 20% [Imaoka *et al*., 2019], the final sample size for power calculation is estimated at 280 participants (140 per group). Power calculations were performed for the primary outcomes, assuming a two-tailed α of 0.05 and a sample size of 140 per group. With these parameters, the study is designed to achieve 80% power to detect an effect size of *d*=0.34, which is an approximately medium effect according to Cohen’s conventional descriptions [Cohen, 1992]. Our analysis will include baseline covariates which increase the power of the study, but for power analysis purposes we ignore these, thereby ensuring that our power estimates are conservative.

The effect size of *d*=0.34 is equivalent to a difference of 0.27 kg/m² in muscle mass index, based on an estimated standard deviation of approximately 0.8 kg/m² [Kwon *et al*., 2015; Fukuoka *et al*., 2019]. Additionally, this effect size represents a difference of 10 seconds in TMT-A and 31 seconds in TMT-B, based on standard deviations of 27.5 seconds and 92.0 seconds, respectively [Hashimoto *et al*., 2006]. Prior studies have demonstrated increases in skeletal muscle mass of 1 kg/m² associated with increased step count [Park *et al*., 2010], improvements of 6.82% (approximately 0.5 kg/m²) in muscle mass with sleep duration exceeding 7 hours [Fu *et al*, 2019], and increases of 0.15 kg/m² with nutrition interventions alone [Strandberg *et al*., 2015]. Furthermore, improvements in cognitive function, as measured by TMT, have shown differences of 10.3 seconds and 11.7 seconds following combined exercise and nutrition interventions, and exercise interventions alone, respectively [Imaoka *et al*., 2019]. Furthermore, studies on cognitive function improvements from nutritional interventions support the validity of our effect size estimation. For example, a trial investigating the effects of Matcha Green Tea Powder reported a significant improvement in MoCA scores in the intervention group (mean change: 1.6, SD: 2.0), with a Cohen’s *d* of 0.50 [Sakurai *et al*., 2020]. While the target effect size of *d*=0.34 in our study is smaller, the observed *d*=0.50 in prior research supports the potential for meaningful effects of nutritional interventions, reinforcing the feasibility of our study design. These findings from existing literature suggest that the calculated effect sizes for our study are appropriate and clinically relevant.

### 2.9 Statistical analysis

Statistical analyses were performed using Bell Curve for Excel version 4.07 (Social Survey Research Information Co. Ltd., Tokyo, Japan) and Python (version 3.12.1; Python Software Foundation, https://www.python.org/) with the following libraries: NumPy (version 1.26.1) for numerical computations, pandas (version 2.2.1) for data manipulation, and SciPy (version 1.14.1) for statistical tests. Visualization was carried out using Matplotlib (version 3.9.2) and Seaborn (version 0.13.2). The significance level was set at *p*<0.05 for all tests, and all tests were two-tailed. Descriptive statistics were calculated for all variables, with continuous variables presented as means and standard deviations, and categorical variables as frequencies and percentages. The primary analysis was conducted based on the intention-to-treat (ITT) principle, including all participants as randomized, regardless of adherence or completion of the protocol. A per-protocol (PP) analysis was performed as a sensitivity analysis, including only participants who completed all study procedures. The ITT analysis included all participants who received either the intervention or the control, regardless of whether they completed the study procedures. This dual analytical approach ensured a comprehensive evaluation of the intervention effects while accounting for potential biases associated with missing data [McCoy, 2017].

P-values for comparisons of baseline and follow-up data between the intervention and control groups were calculated using independent t-tests for continuous variables and chi-square tests for categorical variables. To assess the intervention effects on distinct functional domains, analysis of covariance (ANCOVA) was conducted separately for general cognitive function (MoCA), muscle mass, and physical frailty (Fried frailty phenotype index), with changes in scores from pre-to post-intervention as the dependent variable and group assignment (intervention vs. control) as the independent variable. For executive function, which was assessed using two related measures (TMT-A and TMT-B), ANCOVA was performed with Benjamini-Hochberg correction for multiple comparisons.

The selection of covariates was based on established factors known to influence cognitive function and physical performance in older adults. Age was included as it is strongly associated with cognitive decline and muscle mass loss [Sui *et al*., 2020]. Sex was controlled due to documented differences in cognitive aging patterns and muscle mass between males and females [McCarrey *et al*., 2016; Schorr *et al*., 2018]. Years of education was included as it is a well-established protective factor against cognitive decline and has been consistently associated with cognitive reserve [Stern, 2012]. BMI was controlled for given its known relationship with both cognitive function and physical frailty [García-Ptacek *et al*., 2014; Hubbard *et al*., 2010]. ApoE status (presence of ε4 and ε2 alleles) was included as a covariate due to its significant role in cognitive aging, with the ε4 allele being a major genetic risk factor for cognitive decline, while the ε2 allele is associated with protective effects [Liu *et al*., 2013]. Baseline scores for each outcome measure were included as covariates to control for initial differences between groups, following standard practices in intervention research [van Breukelen, 2006]. This comprehensive set of covariates allows for a robust evaluation of the intervention effects while accounting for key demographic, anthropometric, genetic, and baseline performance factors that could influence the outcomes.

A subgroup analysis was performed on participants who expressed willingness to keep food records at the start of the study and receive nutritional alerts, comparing their outcomes to those in the control group who used the Asken app for dietary tracking but did not receive intervention alerts. The same ANCOVA approach was applied, with changes in scores from pre-to post-intervention as the dependent variable, group assignment as the independent variable, and baseline scores, age, sex, years of education, BMI, and ApoE status as covariates. For the executive function measures, the Benjamini-Hochberg correction was similarly applied to account for multiple comparisons.

The ITT analysis addressed missing data in the primary outcomes (MoCA, Frailty Index, TMT-A, TMT-B, and muscle mass) through multiple imputation using the iterative chained equations (ICE) method. This approach minimized bias and preserved statistical power by estimating plausible values for missing data [Sterne *et al*., 2009]. The scikit-learn library’s IterativeImputer module was employed to perform the imputations, iteratively refining missing value estimates until convergence was achieved, with a maximum of 50 iterations allowed per imputation. Convergence stability was validated to ensure the accuracy of the imputations. The imputation model included baseline and follow-up values for each outcome, as well as predictors selected based on their relevance to the study outcomes. For cognitive measures (MoCA and TMT), predictors included age, sex, years of education, BMI, and ApoE genotype (ε2 and ε4). For the Frailty Index and muscle mass, predictors included age, sex, years of education, body composition metrics (e.g., muscle mass, body fat percentage), ApoE genotype, and the five components of the Frailty Index (weight loss, weakness, exhaustion, slowness, and low physical activity).

### 2.10 Safety Assessment and Adverse Event Tracking

The safety of participants was continuously monitored throughout the study. At follow-up visit, participants were systematically questioned about any negative experiences or unexpected health changes. Additionally, participants were instructed to report any concerns or unusual symptoms to the study team immediately via a dedicated phone line or email address.

All reported events were evaluated by the principal investigator to determine if they were unanticipated and potentially related to the study. Events were classified as adverse events (AEs) or serious adverse events (SAEs). SAEs were defined as any adverse event that resulted in death, was life-threatening, required hospitalization or prolongation of existing hospitalization, resulted in persistent or significant disability/incapacity.

### 2.11 Ethical Considerations

The study was approved by the ethics committee of The University of Tokyo (approval number: 22-360), and registered in the Clinical Trials gov (ClinicalTrials.gov ID: NCT06135740). All participants provided written informed consent. Data privacy and security were ensured through encryption of all personal data and secure storage on password-protected servers. The study is reported according to CONSORT reporting guidelines (https://www.equator-network.org/reporting-guidelines/consort/)

## 3. Results

### 3.1 Primary Analysis

#### 3.1.1 Participant Characteristics

A total of 278 participants completed all study procedures (140 in the intervention group, 138 in the control group). **Figure 1** shows the flow of participants through the study.

**Figure 1:**
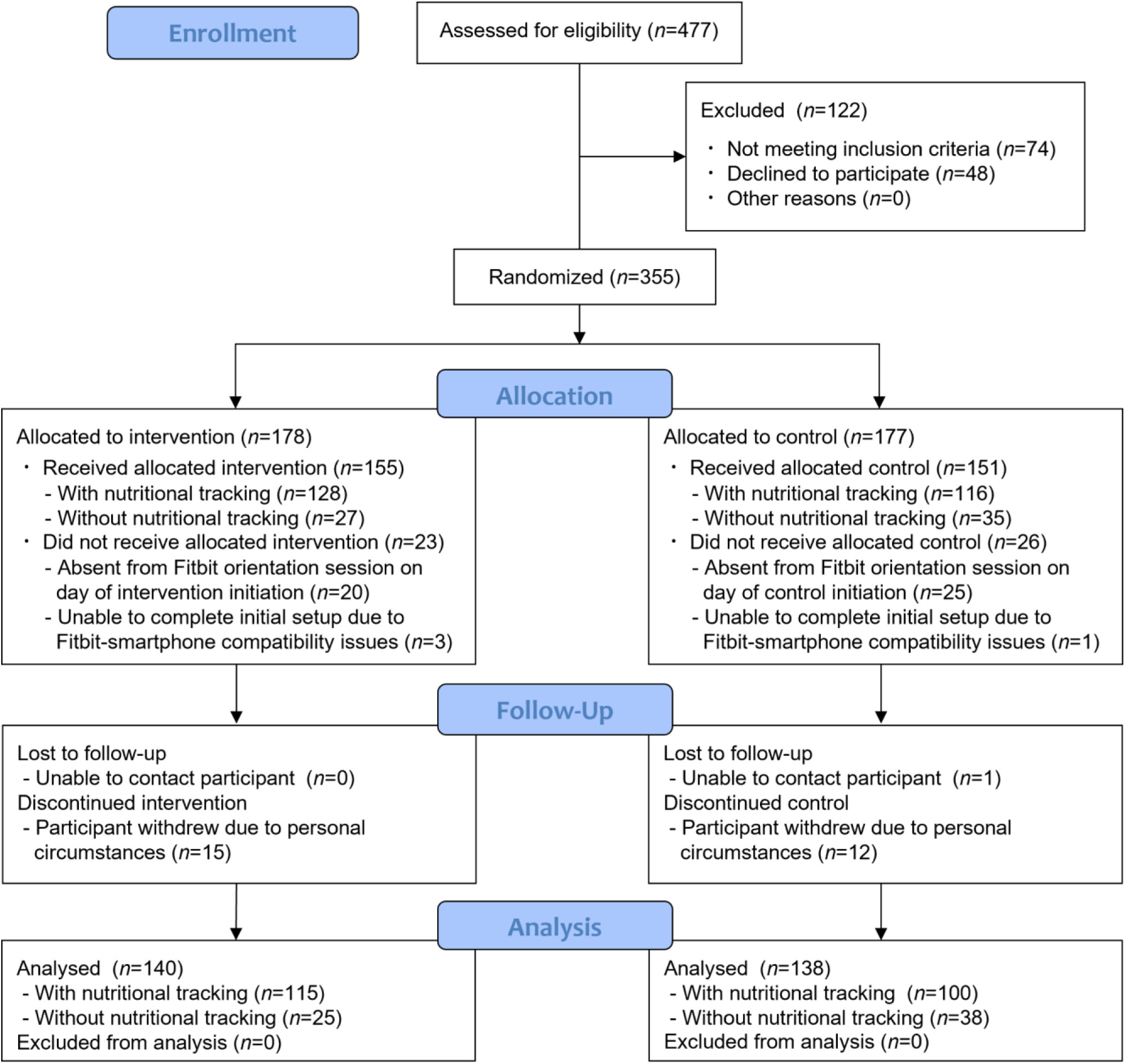
CONSORT diagram of participant enrollment, allocation, follow-up, and analysis.

**Table 3** presents the baseline characteristics of the allocated participants. No significant differences were observed between the groups in most baseline characteristics, including age, sex distribution, years of education, body weight, BMI, body fat percentage, muscle weight, SMI, ApoE4 carrier status, MoCA, TMT-A/B, and Frailty index. However, the proportion of ApoE2 carriers was significantly higher in the control group (*p*=0.044).

**Table 3:** Baseline Characteristics of Allocated Participants.

| Characteristic | Intervention (n=178) | Control (n=177) |
| --- | --- | --- |
| Age (year) | 75.0 ± 5.3 | 75.5 ± 5.1 |
| Sex (Male) | 62 (34.8%) | 60 (33.9%) |
| Education (years) | 14.0 ± 2.3 | 14.1 ± 2.4 |
| Body weight (kg) | 54.8 ± 9.7 | 54.7 ± 10.3 |
| BMI (kg/m<sup>2</sup>) | 21.5 ± 2.9 | 21.7 ± 3.2 |
| Body fat percentage (%) | 24.6 ± 8.4 | 24.8 ± 8.3 |
| Muscle mass (kg) | 38.8 ± 7.3 | 38.6 ± 7.2 |
| SMI (kg/m<sup>2</sup>) | 6.6 ± 1.1 | 6.6 ± 1.1 |
| ApoE 2 carrier | 11 (6.2 %) | 20 (11.3%) |
| ApoE 4 carrier | 28 (15.7 %) | 25 (14.1 %) |
| MoCA | 25.1 ± 2.7 | 25.0 ± 3.2 |
| TMT-A | 77.6 ± 39.3 | 80.2 ± 34.1 |
| TMT-B | 112.2 ± 53.9 | 118.1 ± 58.3 |
| Frailty index | 1.4 ± 1.0 | 1.2 ± 0.8 |

Data are presented as mean ± standard deviation for continuous variables and as number (%) for categorical variables. P-values were calculated using independent t-tests for continuous variables and chi-square tests for categorical variables. BMI = Body mass index; SMI = Skeletal muscle mass index; ApoE 2 = Apolipoprotein E ε2 allele. ApoE 4 = Apolipoprotein E ε4 allele.

#### 3.1.2 General Cognitive Function

The intervention group showed significant improvement in cognitive function as measured by the MoCA. The change in total MoCA score from baseline to follow up at 6 months was significantly greater in the intervention group compared to the control group (mean difference: 1.0 (95% CI:0.6 to 1.3) vs 0.2 (−0.2 to 0.5), *p*=0.011; **Table 4**).

**Table 4:** Changes in Primary Outcomes from Baseline to Follow-Up.

| Measures | Baseline | | | Follow-Up | | | $\Delta$ | | |
| --- | --- | --- | --- | --- | --- | --- | --- | --- | --- |
|  | Intervention (n=155) | Control (n=151) | p-value | Intervention (n=155) | Control (n=151) | p-value | Intervention (n=155) | Control (n=151) | p-value |
| MoCA | 25.2 ± 2.6 | 25.5 ± 2.8 | 0.305 | 26.2 ± 2.9 | 25.7 ± 2.8 | 0.152 | 1.0 (0.6 to 1.3) | 0.2 (-0.2 to 0.5) | 0.0004 |
| Muscle mass (kg) | 38.9 ± 7.3 | 38.9 ± 7.5 | 0.953 | 39.2 ± 7.3 | 39.0 ± 7.3 | 0.832 | 0.3 (0.1 to 0.4) | 0.1 (0.0 to 0.3) | 0.290 |
| Frailty index | 1.3 ± 0.9 | 1.2 ± 0.8 | 0.353 | 1.0 ± 0.9 | 1.1 ± 0.8 | 0.148 | -0.3 (-0.5 to -0.2) | -0.1 (-0.2 to 0.1) | 0.029 |

Data are presented as mean ± standard deviation for baseline and follow-up values, and as mean differences with 95% confidence intervals (CI) for changes from baseline to follow-up. P-values were calculated using independent t-tests for between-group comparisons at baseline and follow-up, and analysis of covariance (ANCOVA) for changes, with score changes as the dependent variable, group (intervention or control) as the independent variable, and sex, age, years of education, BMI, and ApoE status as covariates. $\Delta$ = Pre-post change.

#### 3.1.3 Executive Function

Changes in executive function, as measured by the TMT-A and TMT-B, did not show statistically significant differences between the intervention and control groups (TMT-A; 5.9 (2.5 to 9.2) vs 5.9 (2.3 to 9.4), *p*=0.815; TMT-B; 4.3 (−2.4 to 11.0) vs −1.7 (−7.7 to 4.3), *p*=0.502). These results are detailed in **Supplementary Table 2**. Due to potential bias stemming from differing conditions during baseline and follow-up assessments, the findings should be interpreted with caution.

#### 3.1.4 Muscle Mass

Muscle mass showed no significant differences between groups at 6-month follow-up (0.3 (0.1 to 0.4) kg vs 0.1 (0.0 to 0.3) kg, *p*=0.290; **Table 4**). Additionally, no significant between-group differences were observed in the changes of body weight, BMI, body fat percentage, or SMI over the study period (**Supplementary Table 2**).

#### 3.1.5 Frailty

The intervention group showed a significant decrease in frailty index compared to the control group (−0.3 (−0.5 to −0.2) vs −0.1 (−0.2 to 0.1), *p*=0.029; **Table 4**), indicating improvement in frailty status.

### 3.2 Subgroup Analysis of participants with nutritional tracking

Within the intervention group using Fitbit devices, a subset of participants voluntarily opted for additional nutritional intervention. This subgroup (*n*=128, 82.6% of the intervention group) chose to use the “Asken” food management app (Iizuka *et al*., 2022) for dietary tracking. Based on their app-recorded dietary data, these participants received personalized alerts regarding their protein intake. In contrast, the control subgroup (*n*=116, 76.8% of the control group) used the Asken app for dietary tracking but did not receive personalized alerts and protein supplementation. This analysis aimed to evaluate the effects of this comprehensive, opt-in nutritional approach combined with the Fitbit-based intervention. Baseline characteristics were comparable between intervention (*n*=128) and control (*n*=116) groups, except for a significantly higher proportion of ApoE2 carriers in the control group (**Supplementary Table 3**).

This subgroup analysis largely corroborated the findings from the primary analysis, with some notable enhancements in statistical significance. Particularly, the improvement in cognitive function as measured by the MoCA showed a more pronounced effect in the subgroup analysis. Additionally, protein intake, as recorded by the Asken app, showed a significant increase in the intervention subgroup compared to the control subgroup (8.2 (6.0 to 10.5) g/day vs 0.5 (−1.8 to 2.8) g/day, *p*<0.001).

The intervention subgroup showed significant improvement in cognitive function as measured by the MoCA. The change in total MoCA score was significantly higher in the intervention subgroup compared to the control subgroup (1.2 (0.8 to 1.6) vs 0.2 (−0.2 to 0.5), *p*=0.0004; **Table 5**).

**Table 5:**
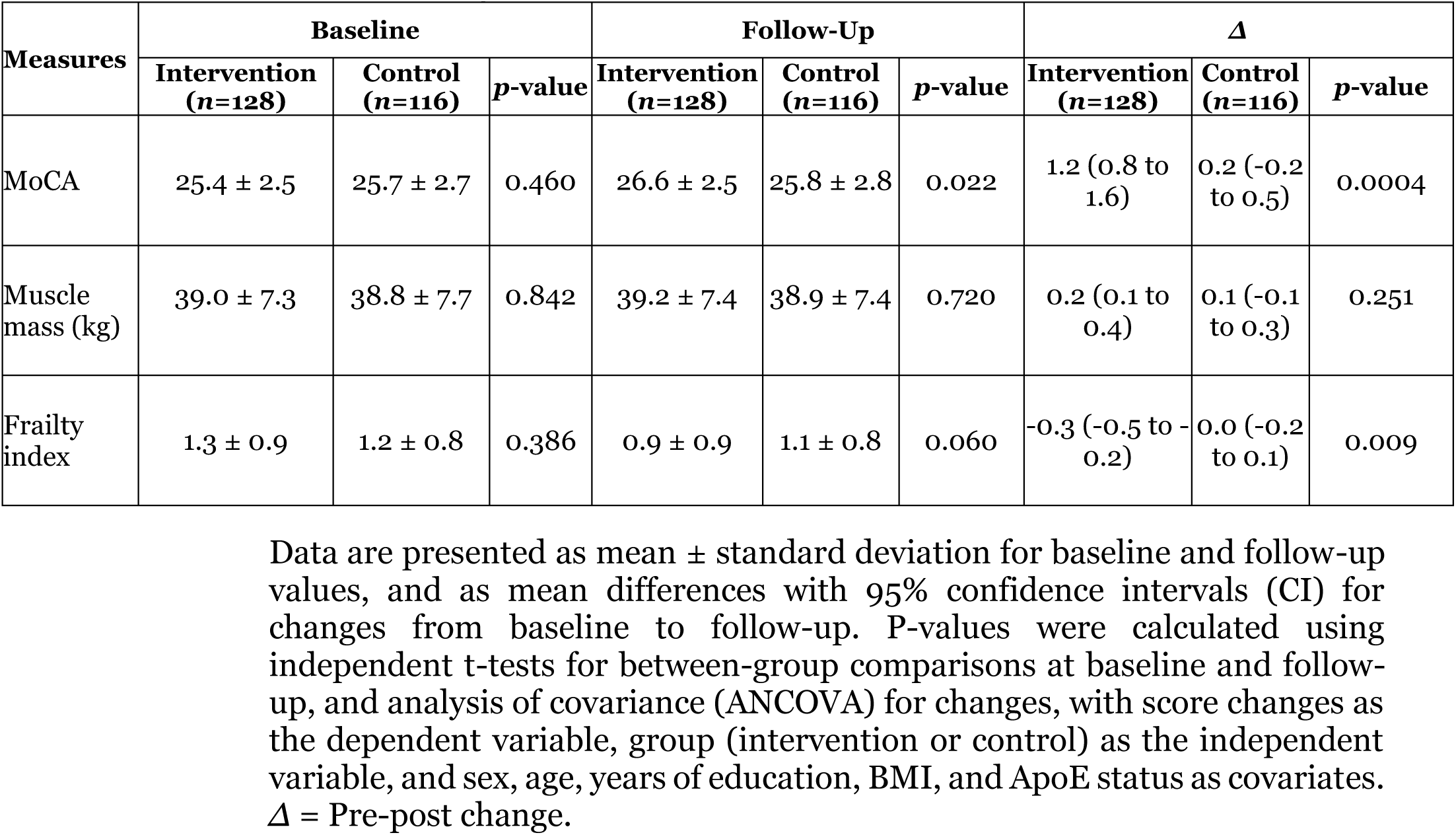
Changes in Primary Outcomes from Baseline to Follow-UP in the participants with nutritional tracking.

| Measures | Baseline | | | Follow-Up | | | $\Delta$ | | |
| --- | --- | --- | --- | --- | --- | --- | --- | --- | --- |
| | Intervention<br>($n=128$) | Control<br>($n=116$) | $p$-value | Intervention<br>($n=128$) | Control<br>($n=116$) | $p$-value | Intervention<br>($n=128$) | Control<br>($n=116$) | $p$-value |
| MoCA | 25.4 ± 2.5 | 25.7 ± 2.7 | 0.460 | 26.6 ± 2.5 | 25.8 ± 2.8 | 0.022 | 1.2 (0.8 to 1.6) | 0.2 (-0.2 to 0.5) | 0.0004 |
| Muscle mass (kg) | 39.0 ± 7.3 | 38.8 ± 7.7 | 0.842 | 39.2 ± 7.4 | 38.9 ± 7.4 | 0.720 | 0.2 (0.1 to 0.4) | 0.1 (-0.1 to 0.3) | 0.251 |
| Frailty index | 1.3 ± 0.9 | 1.2 ± 0.8 | 0.386 | 0.9 ± 0.9 | 1.1 ± 0.8 | 0.060 | -0.3 (-0.5 to -0.2) | 0.0 (-0.2 to 0.1) | 0.009 |

Data are presented as mean ± standard deviation for baseline and follow-up values, and as mean differences with 95% confidence intervals (CI) for changes from baseline to follow-up. P-values were calculated using independent t-tests for between-group comparisons at baseline and follow-up, and analysis of covariance (ANCOVA) for changes, with score changes as the dependent variable, group (intervention or control) as the independent variable, and sex, age, years of education, BMI, and ApoE status as covariates. $\Delta$ = Pre-post change.

Executive function changes, evaluated using the TMT-A and TMT-B, showed no statistically significant differences between the intervention and control subgroups (TMT-A; 6.5 (2.7 to 10.3) vs 5.9 (1.7 to 10.2), *p*=0.815; TMT-B; 3.4 (−4.2 to 11.1) vs −0.9 (−8.0 to 6.1), *p*=0.502; **Supplementary table 4**). Similarly, no significant differences were observed in changes to Muscle mass or other body composition measures (Muscle mass; 0.2 (0.1 to 0.4) vs 0.1 (−0.1 to 0.3), *p*=0.251; **Table 5**) and the intervention subgroup showed a significant improvement in frailty status compared to the control subgroup (−0.3 (−0.5 to −0.2) vs 0.0 (−0.2 to 0.1), *p*=0.009; **Table 5**).

In the subgroup analysis of participants who did not receive the nutritional tracking, no statistically significant differences were observed between the intervention and control groups across all primary outcomes. Changes in cognitive function, frailty status, and other health-related metrics were minimal and comparable between the two groups.

### 3.3 Sensitivity Analysis

Sensitivity analysis using the PP approach confirmed the robustness of the primary findings derived from the ITT analysis. The PP analysis demonstrated significant improvements in the intervention group for MoCA (1.0 (0.6 to 1.4) vs 0.2 (−0.2 to 0.6), *p*=0.021) and the frailty index (−0.3 (−0.5 to −0.2) vs −0.1 (−0.2 to 0.1), *p*=0.040), consistent with the PP analysis. No significant differences were observed between groups for TMT-A and TMT-B (TMT-A; 5.8 (1.7 to 9.8) vs 6.2 (2.1 to 10.4), *p*=0.793; TMT-B; 3.5 (−4.6 to 11.6) vs −1.0 (−8.1 to 6.0), *p*=0.793) or muscle mass (0.0 (0.0 to 0.1) vs 0.0 (0.0 to 0.1), *p*=0.954).

Similarly, in the nutritional tracking subgroup analysis, the PP results mirrored the ITT findings, with significant improvements in MoCA (1.2 (0.8 to 1.6) vs 0.2 (−0.3 to 0.6), *p*=0.001) and the frailty index (−0.3 (−0.5 to −0.2) vs −0.1(−0.2 to 0.1), *p*=0.014) in the intervention group, but no significant differences for TMT-A and TMT-B (TMT-A; 8.6 (−2.5 to 19.7) vs 6.2 (−0.1 to 12.4), *p*=0.817; TMT-B; 7.0 (−14.3 to 28.2) vs −4.5 (−16.7 to 7.7), *p*=0.817) or muscle mass (0.0 (0.0 to 0.1) vs 0.0 (0.0 to 0.1), *p*=0.928).

### 3.4 Adverse Events and Technology Usability

No serious adverse events related to the intervention were reported throughout the study period. However, 10.7% of participants in the intervention group reported experiencing mild skin irritation, specifically itching on the wrist, attributed to the Fitbit watch band, while 21.1% of participants in the control group reported similar symptoms. Notably, the control group participants wore the Fitbit only during the first and last two weeks of the study. This limited wearing period may have contributed to the higher rate of reported irritation, as participants lacked time to adjust to the device. To address this issue, participants experiencing discomfort were offered the option to exchange their silicone watch band for a fabric alternative at no cost, ensuring participant comfort and adherence to the study protocol.

The mean SUS score for the Fitbit device in the intervention group was 57.0 ± 11.5, suggesting usability that Bangor *et al*. (2009) would characterize as “OK.”

## 4. Discussion

### 4.1 Main Findings

This 6-month randomized controlled trial provides evidence that personalized, multifaceted interventions delivered by Fitbit prompts delivered to higher risk community-dwelling elderly adults in Japan was associated with small but statistically significant increases in overall cognitive function, reduction in frailty and improved physical health outcomes over 6 months. The use of personalized lifestyle notifications underscores the potential importance of tailored digital health interventions, rather than a ‘one size fits all’ approach in promoting wellness. By leveraging real-time data from wearable devices, we were able to provide timely, relevant advice to participants.

We found that general cognitive function, as measured by the MoCA, increased by 1.0 in the intervention group, which is considered a meaningful difference [Lindvall *et al*., 2024], compared to 0.2 in the control group. This finding aligns with previous research suggesting that multicomponent interventions can effectively maintain or enhance cognitive function in older adults [Rosenberg *et al*., 2018].

A second key finding from this trial was a significant reduction in frailty scores in the intervention group, with frailty score changing by −0.2 between intervention and control groups at follow up. Frailty is a critical health concern in older adults, associated with increased risk of adverse health outcomes [Fried *et al*., 2001; Hoogendijk *et al*., 2019]. Our results suggest that wearable technology-based interventions could play a role in frailty prevention and management, potentially through promoting physical activity and better diet and sleep patterns. This aligns with previous research indicating that lifestyle interventions can modify frailty status [Dent *et al*., 2019].

The subgroup analysis of participants who opted for comprehensive nutritional tracking and alert intervention reveals several important findings that warrant detailed discussion. The most notable outcome was the enhanced improvement in cognitive function among participants who received both Fitbit-based activity monitoring and nutritional tracking, as evidenced by the significantly greater increase in MoCA scores compared to the control group (1.1 (0.7 to 1.5) vs 0.2 (−0.2 to 0.5), *p*=0.0004). This finding aligns with previous research by Ngandu et al. (2015), which demonstrated that combined lifestyle interventions targeting both physical activity and nutrition tend to yield superior cognitive outcomes compared to single-domain interventions.

Given that technology adoption can be a barrier in this population [Kononova *et al*., 2019], our findings of a high completion rate (90.3%) among participants who received the intervention, coupled with the SUS score of 57.0, suggest high acceptance and engagement among our older adult participants. This provides strong support for the feasibility of using wearable technology as a platform for delivering health interventions to older adults, and is consistent with other research that has emphasized that a user-friendly design and straightforward feedback are crucial in encouraging technology use among older adults [Farivar *et al*., 2020].

### 4.2 Comparison with Existing Literature

The observed improvements in cognitive function and frailty status suggest that our comprehensive intervention operated through multiple pathways. Personalized lifestyle notifications were designed to address key health behaviors, including physical activity, sleep patterns, and daily routines, which are known to influence cognitive and physical health. These findings align with prior research showing that improvements in these lifestyle factors can enhance cognitive performance and reduce frailty risk through mechanisms such as improved vascular health and cerebral blood flow, reduced inflammation, and enhanced neuroplasticity [Dedeyne *et al*., 2017; Kivipelto *et al*., 2020; Phillips *et al*., 2018].

The significant improvement in general cognitive function, as measured by MoCA, underscores the effectiveness of the intervention’s multi-domain approach. Notably, the subgroup that received opt-in notifications related to nutrition experienced even greater improvements, highlighting the effectiveness of the intervention’s approach targeting multiple areas, including exercise, sleep, and nutrition. For example, the FINGER trial demonstrated that a 2-year multidomain intervention, incorporating physical activity, cognitive training, and dietary guidance, improved cognitive performance and slowed decline in older adults at risk for dementia [Ngandu *et al*., 2015; Rosenberg *et al*., 2018].

The primary focus of the nutritional intervention in this study was the delivery of tailored notifications encouraging improved nutritional status through increased protein intake. Protein balanced food products were offered as an optional support to address financial barriers, providing a convenient way for some participants to boost their protein intake. While previous research by van de Rest et al. (2014) showed that 30g of daily protein supplementation improved cognitive functions in frail older adults, the average increase in protein intake in our study was approximately 8.2g per day. This suggests that the observed improvements in cognitive and physical function cannot be solely attributed to protein supplementation.

The protein balanced foods, comprising equal parts animal (chicken) and plant-based (soy) protein, may have supported dietary changes. Chicken breast is rich in imidazole dipeptides like carnosine and anserine, known for their antioxidant, anti-inflammatory, and neuroprotective properties [Kaneko et al., 2017; Lei et al., 2024; Masuoka et al., 2021], while soy proteins are high in isoflavones and other bioactive compounds associated with cognitive benefits in older adults [Soni et al., 2014]. However, given the modest protein increase and the multifaceted nature of the intervention, the observed benefits are more likely due to a combination of improved dietary habits, physical activity monitoring, and personalized guidance from notifications.

Previous studies have demonstrated that wearable device-based interventions effectively improve health behaviors among older adults, particularly when combined with behavioral strategies. Lyons *et al*. [2017] reported that an intervention using a wearable activity monitoring system combined with telephone counseling significantly increased physical activity levels in older adults. Similarly, Cadmus-Bertram *et al*. [2015] showed that Fitbit-based interventions effectively increased moderate-to-vigorous physical activity in older women through regular feedback and goal-setting approaches. These behavioral changes were found to be sustainable, especially when step count monitoring was combined with personalized goal-setting strategies [McMahon *et al*., 2016]. While these studies established the effectiveness of wearable device interventions in modifying health behaviors, their direct impact on clinical outcomes had not been thoroughly investigated.

Our study builds on these findings by suggesting that the behavioral changes induced by Fitbit-based interventions may have contributed to the observed improvements in clinical outcomes, particularly in cognitive function and frailty status. The improvements in clinical measures are likely mediated by the same behavioral pathways identified in previous research, supporting the potential of such interventions to yield clinically significant benefits in elderly populations.

Regarding muscle mass, our study did not observe significant changes. This finding aligns with previous research suggesting that factors beyond total protein intake, such as the timing and distribution of intake and the inclusion of resistance exercise, are crucial for muscle protein synthesis [Mamerow *et al*., 2014; Stokes *et al*., 2018]. The lack of a structured resistance exercise component in our intervention may have limited the anabolic response to protein intake, highlighting the importance of combining protein supplementation with resistance training to optimize muscle mass gains.

### 4.3 Strengths and Limitations

Our study had several key strengths. First, we deliberately selected a study population with risk factors for cognitive and physical decline and who had common comorbidities, yet who were still living at home. We believe this supports the external validity of our findings to similar populations and settings. Second, the random allocation to control and intervention arms minimised the potential effect of multiple confounders which would limit the internal validity of study findings. Third, among those who received the intervention or control allocation, dropout rates were low (9.7% and 8.6% respectively), and the effects of the intervention on the primary outcomes were similar when PP vs ITT analyses. Fourth, combined with the low dropout rates and relatively high SUS scores, implies high tolerability for this type of intervention in this population.

We also acknowledge several potential limitations. First, the 6-month duration of our intervention may not capture the long-term impacts on cognitive function and physical health in this population. Longer follow-up periods are crucial to assess the sustainability of the observed improvements and to explore potential impacts on clinical outcomes such as dementia incidence or disability rates. Second, while we controlled for several potential confounding factors, unmeasured variables such as social engagement or cognitive activities outside the intervention could have influenced results. Additionally, it is important to note that protein supplements were only provided in the intervention group, which may have contributed to the observed improvements in cognitive and physical health. Future studies should aim to capture a broader range of lifestyle factors to provide a more comprehensive understanding of the intervention’s effects within the context of participants’ daily lives. Third, although our study population was diverse, it may not be fully representative of the broader older adult population, particularly those with more advanced cognitive impairment or severe frailty, or those without access to smartphones. However, the findings from the ANCOVA, adjusting for covariates such as age, sex, education, body composition, genetic information, and baseline outcome scores, suggest that the intervention had a consistent effect across different participant characteristics. Further research is needed to investigate the generalizability of similar interventions in more vulnerable populations including those with lower technological literacy and access. Fourth, while the control group was provided with Fitbit devices only for baseline activity and sleep measurements, unintended engagement with the Fitbit device by some control group participants may have underestimated the intervention’s true effect size. Finally, while general cognitive function showed significant improvement, the results for executive function measured by the TMT require cautious interpretation. At baseline, the TMT was re-administered on a different day due to a procedural error, and at follow-up, the TMT was conducted alongside other cognitive assessments, potentially causing fatigue and impacting performance. This may explain why both the intervention and control groups showed significantly lower scores at follow-up in the TMT-A, despite no significant differences between groups. These differences in testing conditions likely introduced bias, limiting the reliability of the TMT results.

While we observed significant improvements across several domains, the specific mechanisms driving these changes remain unclear. Future studies should incorporate more detailed physiological and neuroimaging measures to better understand the underlying processes contributing to the observed benefits.

### 4.4 Implications for Researchers, Clinicians and Policy Makers

Looking ahead, several key directions for future research emerge from our findings. Longer-term studies are essential to assess the durability of intervention effects and potential impact on clinical outcomes. Additionally, investigating the effectiveness of similar interventions in more diverse older adult populations, including those with varying levels of cognitive impairment and frailty, will be crucial for broadening the applicability of this approach. Exploring potential synergies between wearable technology-based interventions and other forms of cognitive and physical training could yield even more robust interventions. The development and testing of more advanced personalization algorithms to further tailor interventions to individual needs and preferences also presents an exciting avenue for enhancing the efficacy of such programs. Finally, research that integrates additional sensors and data sources could provide a more comprehensive picture of health status and behavior, potentially leading to more nuanced and effective interventions. As technology continues to advance, the potential for wearable-based interventions to support healthy aging (as well as to provide timely alerts to the emergence of disease) will likely expand, offering new opportunities to promote cognitive health and overall well-being in older adults.

For clinicians and caregivers, our study provides evidence that personalized, Fitbit-based interventions should be part of technology driven approaches to promoting healthy aging. This approach aligns with the growing recognition of the importance of wearable device-based personalized medicine [Cilli *et al*., 2022]. Identifying the participants for whom this type of intervention is most likely to be effective (and those in whom it is not) will be essential for more widespread deployment. Moreover, evidence to support the cost effectiveness of this type of intervention may be vital to encourage policies to support accessibility to this technology among vulnerable individuals across the community. Policies that support adoption and implementation of this type of intervention, combined with evidence for longer term cost effectiveness, will be valuable for such wider use across such communities.

Finally, as our understanding of the interplay between lifestyle factors, technology use, and healthy aging continues to grow, we move closer to developing more effective strategies for promoting health and well-being in our aging societies. The promising results of this study provide a strong foundation for future research and development in this critical area of public health.

## 5. Conclusion

Our study demonstrates that personalized Fitbit-based interventions can effectively enhance cognitive function and reduce frailty progression in older adults. The wearable technology platform proved to be a useful tool for delivering comprehensive health interventions, with its tracking and notification system supporting positive health outcomes, including improvements in both cognitive and physical measures. These findings underscore the potential of digital health solutions in promoting healthy aging, particularly when integrated into a holistic approach to health management. The Fitbit platform’s ability to provide real-time health insights and personalized recommendations appears to be a key driver in supporting behavioral changes that contribute to improved health outcomes. Notably, when combined with optimized protein intake, participants showed enhanced benefits, suggesting the value of comprehensive lifestyle management that includes both digital monitoring and attention to key nutritional factors.

However, it is important to note that while our intervention showed significant benefits, further research is needed to optimize the implementation of wearable technology across diverse older adult populations. Future studies should focus on refining these interventions, exploring their long-term impacts, and investigating the specific mechanisms through which digital health monitoring contributes to improved health outcomes. As our understanding of the relationship between technology-enabled lifestyle monitoring and healthy aging continues to grow, we move closer to developing more effective strategies for promoting health and well-being in our aging societies. The promising results of this study provide a strong foundation for future research and development in this critical area of public health.

## 8. Author Contributions

Conceptualization, K.S., Y.S., J.L., S.W., and T.H.; Methodology, K.S., Y.S., J.L., and T.H.; Formal Analysis, K.S., I.S., H.K. S.A., R.T., M.W., M.F., and T.H.; Investigation, K.S., I.S., H.K., S.A., R.T., M.W., M.F., S.S., T.G., R.K., K.M., T.K., and T.H.; Resources, N.I., K.K., S.K., N.K., T.M., T.T., T.I., K.K., Y.S., H.K., J.L., and T.H.; Data Curation, K.S., I.S., H.K., S.A., R.T., M.W., M.F., S.S., T.G., R.K., and T.H.; Writing—original draft preparation, K.S. and T.H.; Writing—review and editing, I.S., H.K., R.T., M.W., M.F., S.S., T.N., T.M., K.K., Y.S., M.T., J.L, S.W., and T.H.; Visualization, K.S., I.S., and H.K.; Supervision, T.H.; Project Administration, Y.S., H.K., J.L., and T.H. All authors have read and agreed to the published version of the manuscript.

## 10. Institutional Review Board Statement

The study was conducted in accordance with the Declaration of Helsinki, and approved by the Ethics Committee of the University of Tokyo (approval number: 22-360), and registered in the Clinical Trials gov (ClinicalTrials.gov ID: NCT06135740).

## 11. Informed Consent Statement

Written informed consent was obtained from all subjects involved in the study.

## 12. Data Availability Statement

Data supporting the results of this study are available from the corresponding author upon reasonable request.

## Acknowledgments

The authors are grateful to the staff of the Community Health Promotion Laboratory for their assistance with data collection.

---

**Supplementary Table 1:** Nutritional Composition and Ingredients of Protein Balanced Food.

| Nutrient | Amount per 100g | Ingredients |
| --- | --- | --- |
| Energy (kcal) | 229.0 | chicken, granular soybean protein, vegetable oil, onion, breadcrumbs, bonito seasoning, kelp extract, ginger, reduced starch syrup, salt, soy sauce, refined sugar, frying oil, seasoning (amino acids, etc.), methylcellulose, acidulant, xylose, flavoring, phosphate |
| Protein (g) | 17.1 |  |
| Fat (g) | 15.0 |  |
| Carbohydrates (g) | 8.3 |  |
| - Glucide (g) | 4.5 |  |
| - Dietary Fiber (g) | 3.8 |  |
| Sodium (mg) | 565.0 |  |
| Salt Equivalent (g) | 1.4 |  |
| Water Content (g) | 57.4 |  |
| Ash Content (g) | 2.2 |  |
| Anserine (mg) | 179.6 |  |
| Carnosine (mg) | 55.9 |  |

Nutritional composition of the protein balanced food is presented per 100 grams, including macronutrients, minerals, and bioactive compounds. Carbohydrates are further divided into glucide and dietary fiber. Values are expressed in kilocalories (kcal), grams (g), or milligrams (mg) as appropriate, and the full list of ingredients utilized in its preparation.

**Supplementary table 2:** Changes in TMT scores and Body Composition Measures from Baseline to Follow-Up.

| Measures | Baseline | | | Follow-Up | | | $\Delta$ | | |
| --- | --- | --- | --- | --- | --- | --- | --- | --- | --- |
|  | Intervention (n=155) | Control (n=151) | p-value | Intervention (n=155) | Control (n=151) | p-value | Intervention (n=155) | Control (n=151) | p-value |
| TMT-A | 85.8 ± 29.7 | 87.1 ± 27.0 | 0.669 | 91.6 ± 29.5 | 93.0 ± 28.8 | 0.683 | 5.9 (2.5 to 9.2) | 5.9 (2.3 to 9.4) | 0.815 |
| TMT-B | 125.0 ± 39.9 | 127.9 ± 49.5 | 0.578 | 129.3 ± 49.5 | 126.2 ± 47.3 | 0.571 | 4.3 (-2.4 to 11.0) | -1.7 (-7.7 to 4.3) | 0.502 |
| Body weight (kg) | 55.1 ± 9.6 | 55.0 ± 10.5 | 0.927 | 55.1 ± 9.2 | 55.1 ± 10.2 | 0.966 | 0.0 (-0.3 to 0.2) | 0.1 (-0.1 to 0.4) | 0.410 |
| BMI (kg/m<sup>2</sup>) | 21.7 ± 2.9 | 21.7 ± 3.1 | 0.875 | 21.7 ± 2.8 | 21.8 ± 3.1 | 0.741 | 0.0 (-0.1 to 0.1) | 0.0 (0.1 to 0.1) | 0.379 |
| Body fat percentage (%) | 24.9 ± 8.5 | 24.9 ± 8.1 | 0.999 | 24.5 ± 8.3 | 24.4 ± 7.9 | 0.922 | -0.3 (-0.7 to 0.1) | -0.4 (-0.9 to 0.0) | 0.735 |
| SMI (kg/m<sup>2</sup>) | 6.6 ± 1.1 | 6.6 ± 1.1 | 0.935 | 6.7 ± 1.0 | 6.7 ± 1.1 | 0.900 | 0.0 (0.0 to 0.1) | 0.0 (0.0 to 0.1) | 0.954 |

Data are presented as mean ± standard deviation for baseline and follow-up values, and as mean differences with 95% confidence intervals (CI) for changes from baseline to follow-up. P-values were calculated using independent t-tests for between-group comparisons at baseline and follow-up, and analysis of covariance (ANCOVA) for changes, with score changes as the dependent variable, group (intervention or control) as the independent variable, and sex, age, years of education, BMI, and ApoE status as covariates. Multiple comparisons were adjusted using the Benjamini-Hochberg method. $\Delta$ = Pre-post change.

**Supplementary Table 3:** Baseline Characteristics of Allocated Participants in the Nutritional Tracking Subgroups.

| Characteristic | Intervention (n=128) | Control (n=116) |
| --- | --- | --- |
| Age (year) | 74.2 ± 5.1 | 74.6 ± 4.9 |
| Sex (Male) | 45 (35.2%) | 40 (34.5%) |
| Education (years) | 14.1 ± 2.3 | 14.2 ± 2.4 |
| Body weight (kg) | 55.1 ± 10.0 | 55.0 ± 10.9 |
| BMI (kg/m<sup>2</sup>) | 21.6 ± 3.0 | 21.7 ± 3.2 |
| Body fat percentage (%) | 24.6 ± 8.5 | 24.9 ± 8.0 |
| Muscle mass (kg) | 39.0 ± 7.3 | 38.8 ± 7.7 |
| SMI (kg/m<sup>2</sup>) | 6.6 ± 1.1 | 6.7 ± 1.2 |
| ApoE 2 carrier | 7 (5.5%) | 13 (11.2%) |
| ApoE 4 carrier | 20 (15.6%) | 19 (16.4%) |
| MoCA | 25.4 ± 2.5 | 25.7 ± 2.7 |
| TMT-A | 85.0 ± 30.9 | 84.1 ± 22.5 |
| TMT-B | 124.5 ± 40.8 | 124.3 ± 47.8 |
| Frailty index | 1.3 ± 0.9 | 1.2 ± 0.8 |

Data are presented as mean ± standard deviation for continuous variables and as number (%) for categorical variables. P-values were calculated using independent t-tests for continuous variables and chi-square tests for categorical variables. BMI = Body mass index; SMI = Skeletal muscle mass index; ApoE 2 = Apolipoprotein E ε2 allele. ApoE 4 = Apolipoprotein E ε4 allele.

**Supplementary Table 4:** Changes in TMT scores and Body Composition Measures from Baseline to Follow-Up in the Nutritional Tracking Subgroups.

| Measures | Baseline | | | Follow-Up | | | $\Delta$ | | |
| --- | --- | --- | --- | --- | --- | --- | --- | --- | --- |
|  | Intervention (n=128) | Control (n=116) | p-value | Intervention (n=128) | Control (n=116) | p-value | Intervention (n=128) | Control (n=116) | p-value |
| TMT-A | 85.0 ± 30.9 | 84.1 ± 22.5 | 0.804 | 91.4 ± 30.7 | 90.0 ± 24.6 | 0.700 | 6.5 (2.7 to 10.3) | 5.9 (1.7 to 10.2) | 0.815 |
| TMT-B | 124.5 ± 40.8 | 124.3 ± 47.8 | 0.978 | 127.9 ± 48.2 | 123.4 ± 45.1 | 0.457 | 3.4 (-4.2 to 11.1) | -0.9 (-8.0 to 6.1) | 0.642 |
| Body weight (kg) | 55.1 ± 10.0 | 55.0 ± 10.9 | 0.898 | 55.2 ± 9.6 | 55.0 ± 10.6 | 0.816 | 0.0 (-0.3 to 0.3) | 0.0 (-0.3 to 0.3) | 0.819 |
| BMI (kg/m<sup>2</sup>) | 21.6 ± 3.0 | 21.7 ± 3.2 | 0.864 | 21.7 ± 2.9 | 21.7 ± 3.2 | 0.900 | 0.0 (-0.1 to 0.1) | 0.0 (-0.2 to 0.1) | 0.872 |
| Body fat percentage (%) | 24.6 ± 8.5 | 24.9 ± 8.0 | 0.806 | 24.4 ± 8.3 | 24.4 ± 8.1 | 0.997 | -0.2 (-0.6 to 0.2) | -0.5 (-0.9 to 0.0) | 0.398 |
| SMI (kg/m<sup>2</sup>) | 39.0 ± 7.3 | 38.8 ± 7.7 | 0.842 | 39.7 ± 7.4 | 38.9 ± 7.4 | 0.720 | 0.2 (0.1 to 0.4) | 0.1 (-0.1 to 0.3) | 0.928 |

Data are presented as mean ± standard deviation for baseline and follow-up values, and as mean differences with 95% confidence intervals (CI) for changes from baseline to follow-up. P-values were calculated using independent t-tests for between-group comparisons at baseline and follow-up, and analysis of covariance (ANCOVA) for changes, with score changes as the dependent variable, group (intervention or control) as the independent variable, and sex, age, years of education, BMI, and ApoE status as covariates. Multiple comparisons were adjusted using the Benjamini-Hochberg method. $\Delta$ = Pre-post change.

## References

Bangor A, Kortum P, Miller J. Determining What Individual SUS Scores Mean: Adding an Adjective Rating Scale. J Usability Stud. 2009 May;4(3):114–23.

Brickwood KJ, Watson G, O’Brien J, Williams AD. Consumer-Based Wearable Activity Trackers Increase Physical Activity Participation: Systematic Review and Meta-Analysis. JMIR Mhealth Uhealth. 2019 Apr 12;7(4):e11819.

Cadmus-Bertram LA, Marcus BH, Patterson RE, Parker BA, Morey BL. Randomized Trial of a Fitbit-Based Physical Activity Intervention for Women. Am J Prev Med. 2015 Sep;49(3):414–8.

Cilli E, Ranieri J, Guerra F, Ferri C, Di Giacomo D. Naturalizing digital and quality of life in chronic diseases: Systematic review to research perspective into technological advancing and personalized medicine. Digit Health. 2022 Dec 18;8:20552076221144857.

Cohen J. A power primer. Psychol Bull. 1992 Jul;112(1):155–9.

Dedeyne L, Deschodt M, Verschueren S, Tournoy J, Gielen E. Effects of multi-domain interventions in (pre)frail elderly on frailty, functional, and cognitive status: a systematic review. Clin Interv Aging. 2017 May 24;12:873–896.

Dent E, Morley JE, Cruz-Jentoft AJ, Woodhouse L, Rodríguez-Mañas L, Fried LP, Woo J, Aprahamian I, Sanford A, Lundy J, Landi F, Beilby J, Martin FC, Bauer JM, Ferrucci L, Merchant RA, Dong B, Arai H, Hoogendijk EO, Won CW, Abbatecola A, Cederholm T, Strandberg T, Gutiérrez Robledo LM, Flicker L, Bhasin S, Aubertin-Leheudre M, Bischoff-Ferrari HA, Guralnik JM, Muscedere J, Pahor M, Ruiz J, Negm AM, Reginster JY, Waters DL, Vellas B. Physical Frailty: ICFSR International Clinical Practice Guidelines for Identification and Management. J Nutr Health Aging. 2019;23(9):771–787.

Farivar S, Abouzahra M, Ghasemaghaei M. Wearable device adoption among older adults: A mixed-methods study. Int J Inf Manage. 2020 Dec;55:102209.

Fitbit. Understanding your Active Zone Minutes. Fitbit Help. Available from: https://help.fitbit.com/articles/en_US/Help_article/1379.htm (Accessed 2024/10/18)

Fried LP, Tangen CM, Walston J, Newman AB, Hirsch C, Gottdiener J, Seeman T, Tracy R, Kop WJ, Burke G, McBurnie MA; Cardiovascular Health Study Collaborative Research Group. Frailty in older adults: evidence for a phenotype. J Gerontol A Biol Sci Med Sci. 2001 Mar;56(3):M146–56.

Fu L, Yu X, Zhang W, Han P, Kang L, Ma Y, Jia L, Yu H, Chen X, Hou L, Wang L, Guo Q. The Relationship Between Sleep Duration, Falls, and Muscle Mass: A Cohort Study in an Elderly Chinese Population. Rejuvenation Res. 2019 Oct;22(5):390–398.

Fukuoka Y, Narita T, Fujita H, Morii T, Sato T, Sassa MH, Yamada Y. Importance of physical evaluation using skeletal muscle mass index and body fat percentage to prevent sarcopenia in elderly Japanese diabetes patients. J Diabetes Investig. 2019 Mar;10(2):322–330.

García-Ptacek S, Faxén-Irving G, Cermáková P, Eriksdotter M, Religa D. Body mass index in dementia. Eur J Clin Nutr. 2014 Nov;68(11):1204–9.

Hamido S, Gu X. The impact of activity sensors on attitudes and behavior among young and elderly people. 2021 IEEE 3rd Global Conference on Life Sciences and Technologies (LifeTech); 2021 Mar 9-11; Nara, Japan. Piscataway (NJ): IEEE; 2021. p. 253-255.

Hashimoto R, Meguro K, Lee E, Kasai M, Ishii H, Yamaguchi S. Effect of age and education on the Trail Making Test and determination of normative data for Japanese elderly people: the Tajiri Project. Psychiatry Clin Neurosci. 2006 Aug;60(4):422–8.

Hoogendijk EO, Afilalo J, Ensrud KE, Kowal P, Onder G, Fried LP. Frailty: implications for clinical practice and public health. Lancet. 2019 Oct 12;394(10206):1365–1375.

Hubbard RE, Lang IA, Llewellyn DJ, Rockwood K. Frailty, body mass index, and abdominal obesity in older people. J Gerontol A Biol Sci Med Sci. 2010 Apr;65(4):377–81.

Iizuka K, Ishihara T, Watanabe M, Ito A, Sarai M, Miyahara R, Suzuki A, Saitoh E, Sasaki H. Nutritional Assessment of Hospital Meals by Food-Recording Applications. Nutrients. 2022 Sep 11;14(18):3754.

Imaoka M, Nakao H, Nakamura M, Tazaki F, Maebuchi M, Ibuki M, Takeda M. Effect of Multicomponent Exercise and Nutrition Support on the Cognitive Function of Older Adults: A Randomized Controlled Trial. Clin Interv Aging. 2019 Dec 11;14:2145–2153.

Kaneko J, Enya A, Enomoto K, Ding Q, Hisatsune T. Anserine (beta-alanyl-3-methyl-L-histidine) improves neurovascular-unit dysfunction and spatial memory in aged AβPPswe/PSEN1dE9 Alzheimer’s-model mice. Sci Rep. 2017 Oct 3;7(1):12571.

Kivipelto M, Mangialasche F, Snyder HM, Allegri R, Andrieu S, Arai H, Baker L, Belleville S, Brodaty H, Brucki SM, Calandri I, Caramelli P, Chen C, Chertkow H, Chew E, Choi SH, Chowdhary N, Crivelli L, Torre R, Du Y, Dua T, Espeland M, Feldman HH, Hartmanis M, Hartmann T, Heffernan M, Henry CJ, Hong CH, Håkansson K, Iwatsubo T, Jeong JH, Jimenez-Maggiora G, Koo EH, Launer LJ, Lehtisalo J, Lopera F, Martínez-Lage P, Martins R, Middleton L, Molinuevo JL, Montero-Odasso M, Moon SY, Morales-Pérez K, Nitrini R, Nygaard HB, Park YK, Peltonen M, Qiu C, Quiroz YT, Raman R, Rao N, Ravindranath V, Rosenberg A, Sakurai T, Salinas RM, Scheltens P, Sevlever G, Soininen H, Sosa AL, Suemoto CK, Tainta-Cuezva M, Velilla L, Wang Y, Whitmer R, Xu X, Bain LJ, Solomon A, Ngandu T, Carrillo MC. World-Wide FINGERS Network: A global approach to risk reduction and prevention of dementia. Alzheimers Dement. 2020 Jul;16(7):1078–94.

Kononova A, Li L, Kamp K, Bowen M, Rikard RV, Cotten S, Peng W. The Use of Wearable Activity Trackers Among Older Adults: Focus Group Study of Tracker Perceptions, Motivators, and Barriers in the Maintenance Stage of Behavior Change. JMIR Mhealth Uhealth. 2019 Apr 5;7(4):e9832.

Kurata H, Meguro S, Abe Y, Sasaki T, Asakura K, Arai Y, Itoh H. Dietary protein intake and all-cause mortality: results from The Kawasaki Aging and Wellbeing Project. BMC Geriatr. 2023 Aug 9;23(1):479.

Kwon HJ, Ha YC, Park HM. The Reference Value of Skeletal Muscle Mass Index for Defining the Sarcopenia of Women in Korea. J Bone Metab. 2015 May;22(2):71–5.

Lei C, Zhang B, Yamaguchi J, Tamura R, Cao X, Liu Y, Seki M, Suzuki Y, Suzuki K, Tanida I, Uchiyama Y, Hisatsune T. Impact of astrocytic C3-production on neuronal mitochondrial dysfunction in tauopathy mouse models. bioRxiv. 2024 Nov 12;2024.11.12.622892.

Lindvall E, Abzhandadze T, Quinn TJ, Sunnerhagen KS, Lundström E. Is the difference real, is the difference relevant: the minimal detectable and clinically important changes in the Montreal Cognitive Assessment. Cereb Circ Cogn Behav. 2024 May 3;6:100222.

Liu CC, Liu CC, Kanekiyo T, Xu H, Bu G. Apolipoprotein E and Alzheimer disease: risk, mechanisms and therapy. Nat Rev Neurol. 2013 Feb;9(2):106–18.

Liu CK, Leng X, Hsu FC, Kritchevsky SB, Ding J, Earnest CP, Ferrucci L, Goodpaster BH, Guralnik JM, Lenchik L, Pahor M, Fielding RA. The impact of sarcopenia on a physical activity intervention: the Lifestyle Interventions and Independence for Elders Pilot Study (LIFE-P). J Nutr Health Aging. 2014 Jan;18(1):59–64.

Lyons EJ, Swartz MC, Lewis ZH, Martinez E, Jennings K. Feasibility and Acceptability of a Wearable Technology Physical Activity Intervention With Telephone Counseling for Mid-Aged and Older Adults: A Randomized Controlled Pilot Trial. JMIR Mhealth Uhealth. 2017 Mar 6;5(3):e28.

Mamerow MM, Mettler JA, English KL, Casperson SL, Arentson-Lantz E, Sheffield-Moore M, Layman DK, Paddon-Jones D. Dietary protein distribution positively influences 24-h muscle protein synthesis in healthy adults. J Nutr. 2014 Jun;144(6):876–80.

Masuoka N, Lei C, Li H, Hisatsune T. Influence of Imidazole-Dipeptides on Cognitive Status and Preservation in Elders: A Narrative Review. Nutrients. 2021 Jan 27;13(2):397.

McCarrey AC, An Y, Kitner-Triolo MH, Ferrucci L, Resnick SM. Sex differences in cognitive trajectories in clinically normal older adults. Psychol Aging. 2016 Mar;31(2):166–75.

McCoy CE. Understanding the Intention-to-treat Principle in Randomized Controlled Trials. West J Emerg Med. 2017 Oct;18(6):1075–1078.

McMahon SK, Lewis B, Oakes M, Guan W, Wyman JF, Rothman AJ. Older Adults’ Experiences Using a Commercially Available Monitor to Self-Track Their Physical Activity. JMIR Mhealth Uhealth. 2016 Apr 13;4(2):e35.

Michie S, Yardley L, West R, Patrick K, Greaves F. Developing and Evaluating Digital Interventions to Promote Behavior Change in Health and Health Care: Recommendations Resulting From an International Workshop. J Med Internet Res. 2017 Jun 29;19(6):e232.

Murman DL. The Impact of Age on Cognition. Semin Hear. 2015 Aug;36(3):111–21.

Ngandu T, Lehtisalo J, Solomon A, Levälahti E, Ahtiluoto S, Antikainen R, Bäckman L, Hänninen T, Jula A, Laatikainen T, Lindström J, Mangialasche F, Paajanen T, Pajala S, Peltonen M, Rauramaa R, Stigsdotter-Neely A, Strandberg T, Tuomilehto J, Soininen H, Kivipelto M. A 2 year multidomain intervention of diet, exercise, cognitive training, and vascular risk monitoring versus control to prevent cognitive decline in at-risk elderly people (FINGER): a randomised controlled trial. Lancet. 2015 Jun 6;385(9984):2255-63.

Park H, Park S, Shephard RJ, Aoyagi Y. Yearlong physical activity and sarcopenia in older adults: the Nakanojo Study. Eur J Appl Physiol. 2010 Jul;109(5):953–61. doi: 10.1007/s00421-010-1424-8. Epub 2010 Mar 25. PMID: 20336310.

Phillips C, Fahimi A. Immune and Neuroprotective Effects of Physical Activity on the Brain in Depression. Front Neurosci. 2018 Jul 26;12:498.

Rosenberg A, Ngandu T, Rusanen M, Antikainen R, Bäckman L, Havulinna S, Hänninen T, Laatikainen T, Lehtisalo J, Levälahti E, Lindström J, Paajanen T, Peltonen M, Soininen H, Stigsdotter-Neely A, Strandberg T, Tuomilehto J, Solomon A, Kivipelto M. Multidomain lifestyle intervention benefits a large elderly population at risk for cognitive decline and dementia regardless of baseline characteristics: The FINGER trial. Alzheimers Dement. 2018 Mar;14(3):263–70.

Sakurai K, Shen C, Ezaki Y, Inamura N, Fukushima Y, Masuoka N, Hisatsune T. Effects of Matcha Green Tea Powder on Cognitive Functions of Community-Dwelling Elderly Individuals. Nutrients. 2020 Nov 26;12(12):3639.

Schorr M, Dichtel LE, Gerweck AV, Valera RD, Torriani M, Miller KK, Bredella MA. Sex differences in body composition and association with cardiometabolic risk. Biol Sex Differ. 2018 Jun 27;9(1):28.

Soni M, Rahardjo TB, Soekardi R, Sulistyowati Y, Lestariningsih, Yesufu-Udechuku A, Irsan A, Hogervorst E. Phytoestrogens and cognitive function: a review. Maturitas. 2014 Mar;77(3):209–20.

Strandberg E, Edholm P, Ponsot E, Wåhlin-Larsson B, Hellmén E, Nilsson A, Engfeldt P, Cederholm T, Risérus U, Kadi F. Influence of combined resistance training and healthy diet on muscle mass in healthy elderly women: a randomized controlled trial. J Appl Physiol (1985). 2015 Oct 15;119(8):918-25.

Stern Y. Cognitive reserve in ageing and Alzheimer’s disease. Lancet Neurol. 2012 Nov;11(11):1006–12.

Sterne JA, White IR, Carlin JB, Spratt M, Royston P, Kenward MG, Wood AM, Carpenter JR. Multiple imputation for missing data in epidemiological and clinical research: potential and pitfalls. BMJ. 2009 Jun 29;338:b2393.

Stokes T, Hector AJ, Morton RW, McGlory C, Phillips SM. Recent Perspectives Regarding the Role of Dietary Protein for the Promotion of Muscle Hypertrophy with Resistance Exercise Training. Nutrients. 2018 Feb 7;10(2):180.

Sui SX, Williams LJ, Holloway-Kew KL, Hyde NK, Pasco JA. Skeletal Muscle Health and Cognitive Function: A Narrative Review. Int J Mol Sci. 2020 Dec 29;22(1):255.

Suthutvoravut U, Tanaka T, Takahashi K, Akishita M, Iijima K. Living with Family yet Eating Alone is Associated with Frailty in Community-Dwelling Older Adults: The Kashiwa Study. J Frailty Aging. 2019;8(4):198–204.

Van Breukelen GJ. ANCOVA versus change from baseline: more power in randomized studies, more bias in nonrandomized studies [corrected]. J Clin Epidemiol. 2006 Sep;59(9):920–5.

van de Rest O, van der Zwaluw NL, Tieland M, Adam JJ, Hiddink GJ, van Loon LJ, de Groot LC. Effect of resistance-type exercise training with or without protein supplementation on cognitive functioning in frail and pre-frail elderly: secondary analysis of a randomized, double-blind, placebo-controlled trial. Mech Ageing Dev. 2014 Mar-Apr;136-137:85-93.

World Health Organization. (2015). World Report on Ageing and Health.

Yamamoto S, Yamasaki S, Higuchi S, Kamiya K, Saito H, Saito K, Ogasahara Y, Maekawa E, Konishi M, Kitai T, Iwata K, Jujo K, Wada H, Kasai T, Nagamatsu H, Ozawa T, Izawa K, Aizawa N, Makino A, Oka K, Momomura SI, Kagiyama N, Matsue Y. Prevalence and prognostic impact of cognitive frailty in elderly patients with heart failure: sub-analysis of FRAGILE-HF. ESC Heart Fail. 2022 Jun;9(3):1574–83.

